# Assessment of inter-laboratory differences in SARS-CoV-2 consensus genome assemblies between public health laboratories in Australia

**DOI:** 10.1101/2021.08.19.21262296

**Authors:** Charles S.P. Foster, Sacha Stelzer-Braid, Ira W. Deveson, Rowena A. Bull, Malinna Yeang, Jane Phan-Au, Mariana Ruiz Silva, Sebastiaan J. van Hal, Rebecca J. Rockett, Vitali Sintchenko, Ki Wook Kim, William D. Rawlinson

## Abstract

Whole-genome sequencing of viral isolates is critical for informing transmission patterns and ongoing evolution of pathogens, especially during a pandemic. However, when genomes have low variability in the early stages of a pandemic, the impact of technical and/or sequencing errors increases. We quantitatively assessed inter-laboratory differences in consensus genome assemblies of 72 matched SARS-CoV-2-positive specimens sequenced at different laboratories in Sydney, Australia. Raw sequence data were assembled using two different bioinformatics pipelines in parallel, and resulting consensus genomes were compared to detect laboratory-specific differences. Matched genome sequences were predominantly concordant, with a median pairwise identity of 99.997%. Identified differences were predominantly driven by ambiguous site content. Ignoring these produced differences in only 2.3% (5/216) of pairwise comparisons, each differing by a single nucleotide. Matched samples were assigned the same Pango lineage in 98.2% (212/216) of pairwise comparisons, and were mostly assigned to the same phylogenetic clade. However, epidemiological inference based only on single nucleotide variant distances may lead to significant differences in the number of defined clusters if variant allele frequency thresholds for consensus genome generation differ between laboratories. These results underscore the need for a unified, best-practices approach to bioinformatics between laboratories working on a common outbreak problem.

## Introduction

The first genome sequences for severe acute respiratory syndrome coronavirus 2 (SARS-CoV-2), the causative pathogen of coronavirus disease 2019 (COVID-19), were released in January 2020 (1, 2). Subsequently, routine whole-genome sequencing (WGS) has been adopted globally as infections of SARS-CoV-2 increase exponentially. As of April 2021, more than a million SARS-CoV-2 genome sequences had been deposited in the GISAID data repository (https://www.gisaid.org/).

Genomic data are useful for understanding the origins and spread of emerging pathogens within the related fields of phylodynamics and molecular epidemiology (3), such as during the 2013–2016 West African Ebola epidemic (4). Accordingly, the rapidly accumulating wealth of genomic data available for SARS-CoV-2 is crucial for real-time research into the origins and ongoing evolution of the virus. For example, comparison of SARS-CoV-2 genome sequences isolated from different patients can inform traditional epidemiological methods and allow the reconstruction of transmission chains (5–9).

Additionally, mutations that have accumulated within SARS-CoV-2 genomes over time have enabled successful typing of SARS-CoV-2 into ‘Pango’ lineages according to a hierarchical nomenclature (10). Classifying SARS-CoV-2 isolates into Pango lineages is important for determining whether any diagnostic mutations, especially amino acid replacements, can be linked to increased transmission and/or any differences in disease severity in patients with COVID-19 (11), thereby allowing “variants of concern” to be designated.

Despite the many benefits of genomic epidemiological approaches for tracking the spread of an emerging pathogen, there are many associated challenges. Initially, genome sequences are not derived from a measurably evolving population, and, as a result, they are not sufficiently informative to allow accurate or precise inferences of evolutionary rates and time scales (12). Rates of basecalling errors vary between different sequencing technologies (13), and platform-specific biases can lead to different genomic regions being more or less error prone depending on the choice of technology (14, 15). In the case of SARS-CoV-2, inferred mutations at some genomic sites are problematic because they are highly homoplastic or disproportionately associated with particular sequencing laboratories or geographic locations (15). Potentially incorporating erroneous mutations into genome sequences can introduce errors in response to pandemic virus spread, given the role of diagnostic mutations in informing public health decisions. For example, errors in the genome sequence of a given SARS-CoV-2 isolate could result in a failure to designate the sequence as a variant of concern. This potential failure could, in turn, negatively impact health outcomes for patients when certain mutations are linked to more severe symptoms (16–18), or could increase the chances of an outbreak when a given variant is known to be more transmissible (11).

Even in situations where one can be confident in the reliability of sequencing data, the optimal way to incorporate mutations from a virus sample into a representative consensus genome is uncertain. In the most simple form, a consensus genome represents the most common allele found at each position in an alignment, such as the alignment gained by mapping sequencing reads against a reference genome. Any minor alleles are generally not reflected in the consensus genome. However, in a positive-sense single-stranded RNA virus such as SARS-CoV-2 (2), any inferred minor alleles that are not erroneous in origin (sequencing errors, artifacts of partial RNA degradation) actually represent intra-host diversity. RNA viruses act as a ‘quasispecies’, with a diverse, dynamically evolving population of viruses being found within a given host (19, 20). Host-to-host transmission of SARS-CoV-2 intra-host single-nucleotide variants (iSNVs) can occur, with some iSNVs restricted to different Pango lineages, consistent with iSNVs potentially influencing the epidemiology of the virus (21). In these situations, solely incorporating the most common nucleotide at each genomic site into the consensus genome does not reflect the diversity of the quasispecies within patients. Alternative bioinformatic strategies include representing the diversity of variants at genomic sites in consensus genomes using IUPAC ambiguity codes based on variant allele frequency thresholds (22).

Caution is warranted in determining whether inferred mutations are real, or are the result of sequencing errors or other biases, when using genomic epidemiological approaches. Careful consideration of how the breadth of viral diversity within a patient is represented is also necessary. However, differences in both wet laboratory and bioinformatics approaches can differentially introduce biases and/or differences in sample consensus genomes. Several studies have evaluated the analytical validity of different sequencing technologies and approaches for WGS analysis of SARS-CoV-2 (14, 23).

Nevertheless, far less attention has been paid to assessing the consistency in sequencing between different laboratories. In this study, we compared sequencing results of 72 SARS-CoV-2 isolates carried out in two laboratories in Sydney, New South Wales, Australia, generated using two analogous bioinformatic workflows, by experienced sequencing teams. We investigated differences in the resulting matched consensus genomes, and their downstream consequences on Pango lineage typing and phylogenetic inference.

## Materials and Methods

### SARS-CoV-2 matched samples

Extracts from SARS-CoV-2-positive nasopharyngeal swabs of patients tested at NSW Health Pathology East Serology and Virology Division (SAViD) are routinely sequenced according to established WGS protocols (14). Each SARS-CoV-2-positive extract is also sent to the Institute of Clinical Pathology and Medical Research (ICPMR), NSW Health Pathology-West, NSW, Australia, for WGS according to their established protocols (8).

Accordingly, the same positive sample is sequenced at both SAViD and ICPMR. Here, we compared the resulting consensus sequences of 72 samples sequenced at ICPMR with their corresponding consensus sequences generated within SAViD. We refer to these corresponding samples as ‘matched samples’.

All samples were collected between early March 2020 and late November 2020. For all samples collected after 1/07/2020, both SAViD and ICPMR worked with the same primary extract, but prior to this date ICPMR were sent secondary extractions from the same positive SARS-CoV-2 swab. Clinical specimens were routinely processed at SAViD for diagnostic purposes. Viral sequence data was identified as exempt from ethics by NSW Health as no human isolate or clinical data was analysed specifically for research purposes in this study. All work carried out by ICPMR was done in accordance with governance regulations from the Human Research Ethics Committees of the Western Sydney Local Health District (2020/ETH00287).

Primary extracts were sequenced and assembled into consensus genomes at SAViD (henceforth: Lab1) according to a custom pipeline (see below). Additionally, ICPMR (henceforth: Lab2) provided raw sequencing reads for matched samples, and these were then assembled into consensus genomes at SAViD according to the same pipeline. Finally, primary assemblies from ICPMR for each of these samples, assembled using the bioinformatics pipeline used by ICPMR (8) (see below), were downloaded from GISAID for the purposes of comparison (Supplementary Table S1). Therefore, each isolate was represented in the dataset three times, once for each assembly. This allows three possible pairwise comparisons per isolate.

### Wet-lab Protocols

At Lab1, automated extraction of total nucleic acid extraction from samples was carried out using the Roche MagNA pure 96 with the Roche MagNA Pure DNA and total NA kit. The superscript IV VILO Master Mix (Thermo Fisher) was used for reverse-transcription of RNA extracts. Prepared cDNA was then amplified separately with each of 14 × ∼2.5-kb amplicons tiling the SARS-CoV-2 genome (24) using the Platinum SuperFi Green PCR Mastermix (Thermo Fisher) with 1.5 μL of cDNA per reaction. PCR products were cleaned using AMPure XP beads (0.8X bead ratio), and quantified using PicoGreen dsDNA Assay (Thermo Fisher). All 14x amplicon products from a given sample were then pooled at equal abundance, and prepped for short-read sequencing using the Illumina DNA Prep Kit, according to the manufacturer’s protocol. Samples were multiplexed using Nextera DNA CD Indexes and sequenced using 150 bp paired-end sequencing on an Illumina MiSeq.

At Lab2, all steps up to and including amplicon generation were the same as at Lab1. Library preparation was carried out using an Illumina Nextera XT Kit, followed by sequencing on an Illumina iSeq or MiniSeq (150 cycles).

### Bioinformatics Protocols

Sequencing reads for each sample derived from Lab1, as well as reads for matched samples provided by Lab2, were processed through a custom bioinformatics pipeline. Briefly, raw reads were processed with *fastp v*0.20.1 (25) to remove any residual sequencing adapters, carry out light quality trimming (-q 20), and only retain reads that pass a minimum length filter (-l 50). Clean reads were then mapped to the NCBI RefSeq assembly of SARS-CoV-2 (NC_045512.2) using *bwa mem* v0.7.17-r1188 (26), with unmapped reads discarded, and primer sequences were soft-clipped from the alignment using *ivar trim* v.1.3 (22) discarding reads that were <30 length. Alignments were converted to pileup format using *samtools mpileup* v1.10 (27) without discarding anomalous read pairs (-A), per-base alignment quality disabled (-B), and no minimum PHRED quality for bases (-Q 0). Using the *mpileup* result, variants were called using *ivar variants*, with a minimum quality score threshold filter (-q 20), a minimum depth of 10 (-m 10) and a minimum variant frequency threshold of 0.1 (-t 0.1). Additionally, consensus genomes were assembled using *ivar consensus*, with a minimum variant frequency threshold of 0.9 (-t 0.9), a minimum quality score of 20 (-q 20), and a minimum depth of 10 (-m 10). This frequency threshold allows any potential intrahost diversity, as reflected by multiple alleles per site, to be represented in resulting consensus genomes as IUPAC ambiguity codes. Any inferred indels were manually inspected in *igv* v2.8.13 (28) to see whether they were well supported by the read evidence and did not occur within homopolymeric regions. Spurious indels were ignored in comparative analyses.

Demultiplexed raw sequencing data from Lab2 were quality trimmed using *Trimmomatic* (v0.36, sliding window of 4, minimum read quality score of 20, leading/trailing quality of 5) (29). The taxonomic identification of samples as predominantly ‘*Betacoronavirus*’ was checked with *Centrifuge* v1.0.4 (30) using a database containing human, prokaryotic and viral sequences. Quality controlled reads were then mapped to the SARS-CoV-2 reference genome using *bwa mem* v0.7.17, with unmapped reads discarded, followed by soft-clipping of reads with *ivar trim*, discarding reads <20 length. Consensus genomes were called using *ivar consensus*, with a minimum depth threshold of 10, minimum quality threshold of 20, and minimum frequency threshold of 0.1. Finally, the 5’ and 3’ untranslated regions were masked from consensus genomes.

### Sequence comparisons

Each consensus genome sequence was first individually aligned to the NC_045512.2 reference genome using *mafft* v7.475 (--thread 1 --quiet --add $seqfile $reference) (31), before being combined into a full alignment. Stochastic wet-lab failures (e.g., amplicon drop out) can lead to regions of the genome not having sufficient depth to call a base, resulting in an N being inserted into the consensus genome. While missing regions of genomes because of low coverage can be problematic for phylogenetic or other clustering analyses, here we instead choose to focus on any differences that occur at high-depth, high-quality bases, since these differences potentially represent systematic biases rather than stochastic sequencing dropouts. Therefore, we did not consider differences between sequences where one base was called as an N and the other base was not. Instead, we focused on comparing sites of the genome where bases were confidently called as a ‘standard’ base (‘A’, ‘T’, ‘G’, ‘C’), or as an ‘ambiguous’ base (any of the IUPAC ambiguity codes).

To calculate differences between matched samples, we used a custom program written in python3 (available from https://github.com/charlesfoster/pairwise_comparisons) that sequentially compares the consensus genomes of matched samples and calculates both the total numbers of differences between samples, and reports the differences themselves. The program takes optional command-line arguments to calculate differences using all sites in the alignment, or instead after ignoring (a) runs of ‘N’ or ‘-’ from the ends of consensus sequences (common in primary assemblies and GISAID uploads, respectively), (b) sites where either, or both, sequences being compared have a gap (‘N’ or ‘-’), (c) sites where one sequence has an indel and the other does not, and/or (d) sites where either, or both, sequences have an ambiguous base (as per IUPAC ambiguity codes).

We chose to compare the similarity of sequences after ignoring the effect of stochastic amplicon drop outs (--ignore_gaps). Additionally, we trimmed the alignment to ensure that both sequences being compared were the same length by removing runs of ‘N’ or ‘-’ from the ends of the alignment (--trim_ends), with the exact number of bases that are trimmed depending on each pair of sequences being compared. Manual investigation of the few insertions in samples relative to the reference genome showed that the insertions occurred in error-prone, homopolymeric runs of bases with spurious evidence provided by sequencing reads. Accordingly, we chose to exclude indel sites from further analyses (-- ignore_indels). Implementing different variant frequency thresholds during consensus genome generation can lead to the introduction of ambiguous bases. Therefore, we compared matched samples both with ambiguous bases taken into account, and after ignoring sites in alignments with ambiguous bases (--ignore_ambiguous). We also replicated our analyses after masking sites that are known to be problematic for SARS-CoV-2 sequencing (15).

To compare the effect of any differences on downstream molecular epidemiological inference, we estimated pangolin lineages for each sequence using *pangolin* v2.3.6 (https://github.com/cov-lineages/pangolin), and determined whether matched samples were assigned the same lineage. Another common goal of molecular epidemiological studies is to place sequences under investigation in a phylogenetic tree. When building a tree including the three assemblies per matched samples, we would expect all three matched samples (henceforth, ‘sample triplets’) to group together in a clade. However, differences in sequence content might prevent this from occurring. Therefore, we inferred a tree using maximum likelihood with iqtree2 with branch support estimated with ultrafast bootstraps, rooting on the Wuhan-Hu-1 reference genome (parameters: -o NC_045512.2 - s $alignment -mset HKY,TIM2,GTR -mfreq F -mrate G,R -bb 1000 -nt auto --keep-ident) (32, 33). The input alignment contained 29513 constant sites and 598 distinct site patterns, of which 388 were parsimony-informative. The IQTREE2 command ensures that 99 starting trees are inferred using parsimony, one starting tree is inferred using a neighbour-joining algorithm, and then a candidate set of best trees are optimised with maximum likelihood nearest neighbour interchange. We then used a custom R function to test whether the clade comprising all sample triplets and their most recent ancestor included a total of three tips. We use the number of tips in this clade as a crude proxy for distance between tips in tree space. However, we acknowledge that a lack of divergence between all sequences in the alignment might lead to identical samples from other patients preventing matched samples from forming sequence triplets in our inferred phylogeny. Likewise, a lack of information in multiple sequence alignments might prevent relationships among sequences from being estimated with robust support. These issues are pertinent to the challenges faced early in the investigation of an emerging pathogen.

## Results

### Pairwise differences

There was a strong concordance in consensus sequences of matched samples generated at Lab1 and Lab2, with a median pairwise identity of 99.996% (SD: 0.011) from 216 pairwise sequence comparisons (Supplementary Table S2). Most matched pairs were identical, with the majority of differences driven by the presence of IUPAC ambiguities in one sequence or the other (Figure 1). However, five out of 216 comparisons (comprising three different samples) possessed differences at a site in the genome where both sequences were confidently assigned a standard nucleotide base (A, T, G, C) (Figure 2; Supplementary Table S2). These strongly supported differential base calls did not occur in primer-binding regions or at known problematic sites in the SARS-CoV-2 genome (15).

**Figure 1:**
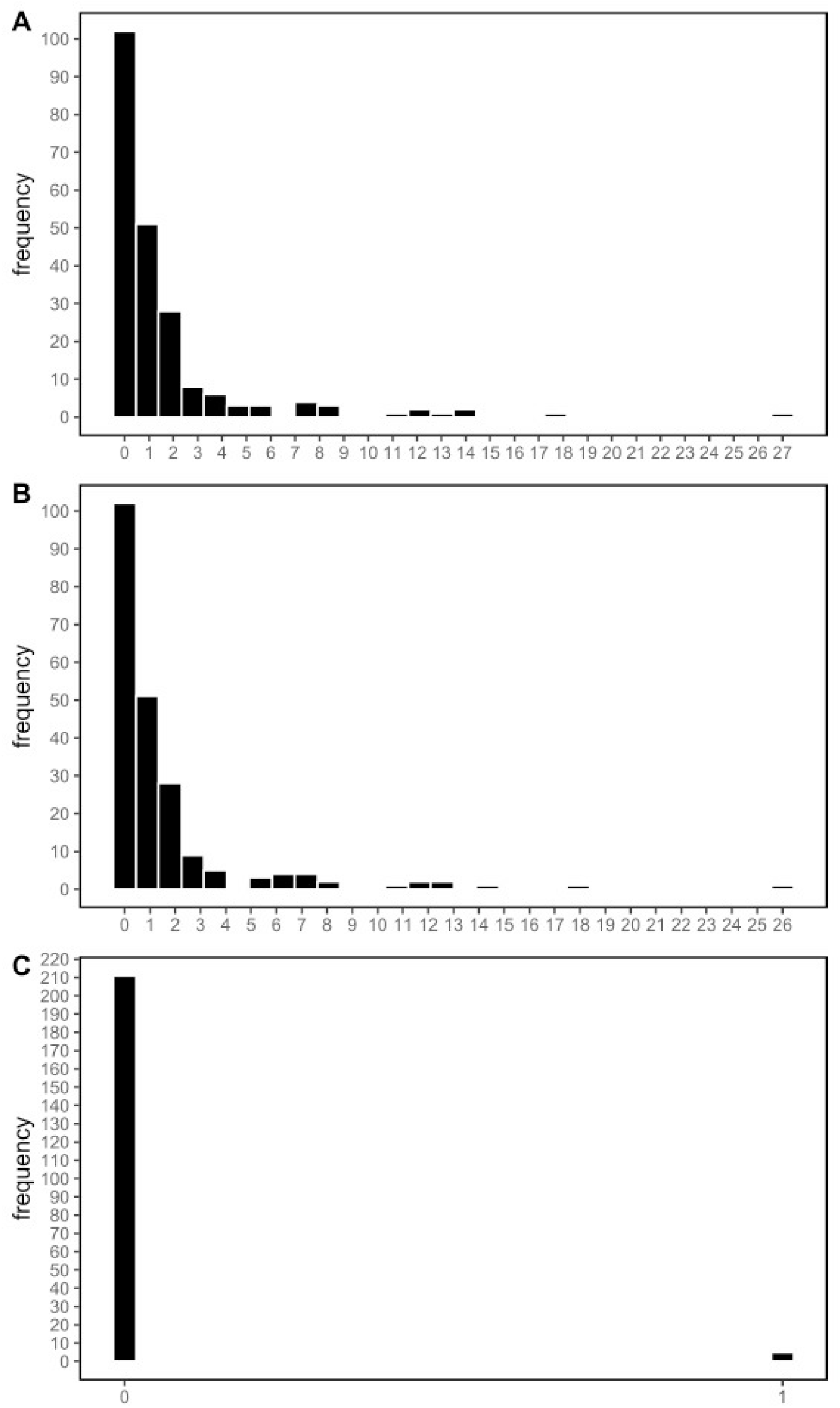
Histograms depicting the frequency of the number of differences between matched SARS-CoV-2 genome sequences. (a) total number of differences; (b) number of differences where either or both sequences being compared had an IUPAC ambiguity code at a given site; (c) number of differences where both sequences being compared had a standard nucleotide base (A, T, G, C) at a given site.

**Figure 2:**
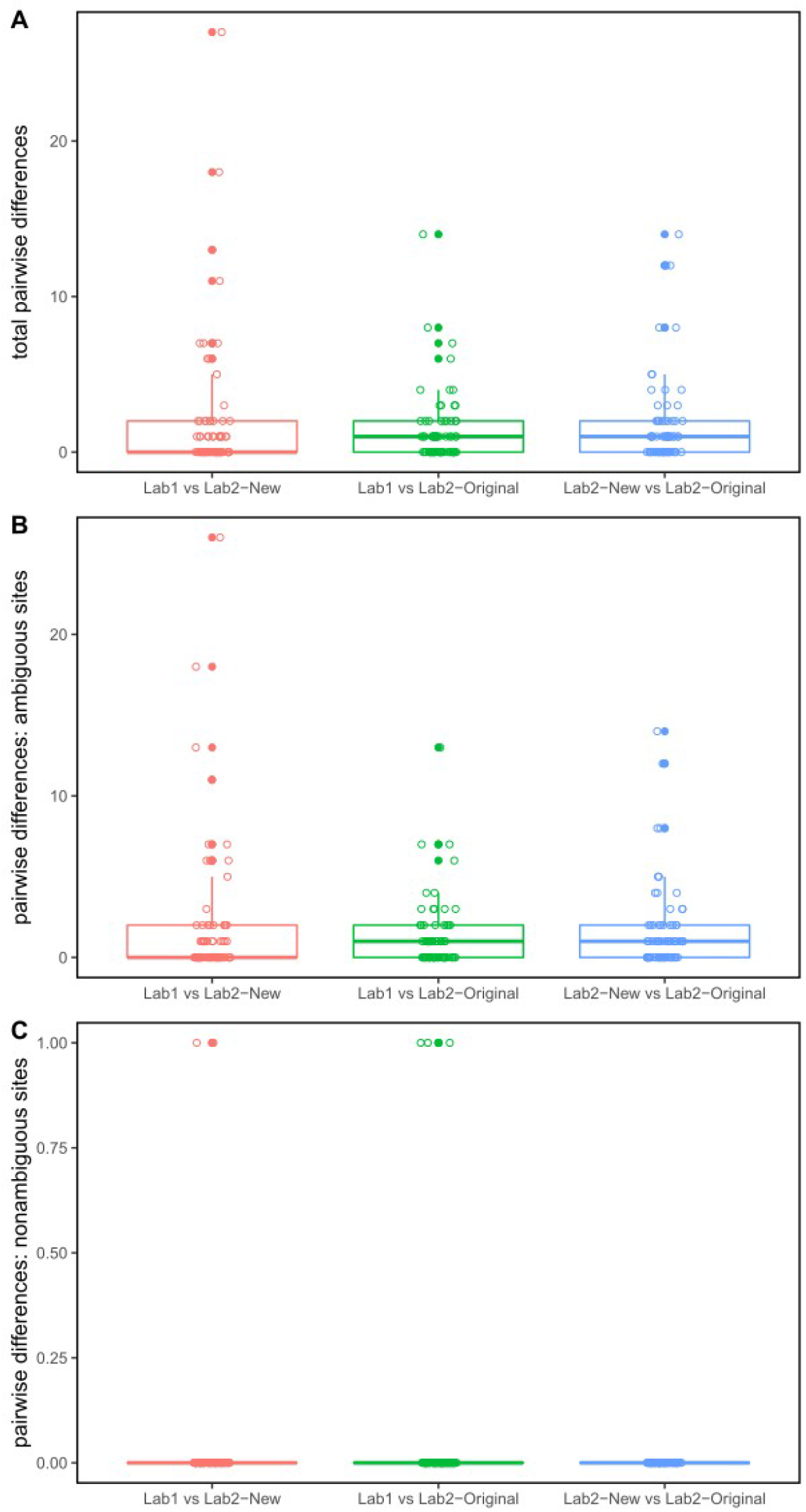
Boxplots depicting the number of differences between matched SARS-CoV-2 genome sequences across 216 pairwise comparisons. (a) total number of differences; (b) number of differences where either or both sequences being compared had an IUPAC ambiguity code at a given site; (c) number of differences where both sequences being compared had a standard nucleotide base (A, T, G, C) at a given site. Within each panel, results are represented based on three classes of analyses: (1) Sequence derived from Lab1 vs sequence derived from Lab2, bothassembled with the bioinformatics protocol of Lab1 (Lab1 vs Lab2−New); (2) Sequence derived from Lab1 assembled with the bioinformatics protocol of Lab1 vs sequence derived from Lab2 assembled with the bioinformatics protocol of Lab2 (Lab1 vs Lab2−Original); (3) Sequence derived from Lab2 assembled with the bioinformatics protocol of Lab1 vs sequence derived from Lab2 assembled with the bioinformatics protocol of Lab2 (Lab2−New vs Lab2−Original).

For further analysis, comparisons between sequences were designated into different ‘classes’:

1. Lab1 vs Lab2-New: Lab1 sample assembled with Lab1 bioinformatics pipeline; Lab2 sample assembled with Lab1 bioinformatics pipeline
2. Lab1 vs Lab2-Original: Lab1 sample assembled with Lab1 bioinformatics pipeline; Lab2 sample assembled with Lab2 bioinformatics pipeline
3. Lab2-New vs Lab2-Original: Lab2 sample assembled with Lab1 bioinformatics pipeline; Lab2 sample assembled with Lab2 bioinformatics pipeline

When investigating the number of differences (SNVs) between each class of sequence pair, the number of differences with and without ambiguous sites included were assessed (Figure 2; Table 1). Comparing the same sequencing reads assembled with different bioinformatics pipelines (class 3: Lab2-New vs Lab2-Original) revealed a number of differences in matched samples when ambiguous sites were included, but, when excluding ambiguous sites, no differences were found. In both other classes of analyses, nearly all differences were able to be attributed to IUPAC ambiguities (Figure 2; Table 1).

**Table 1:**
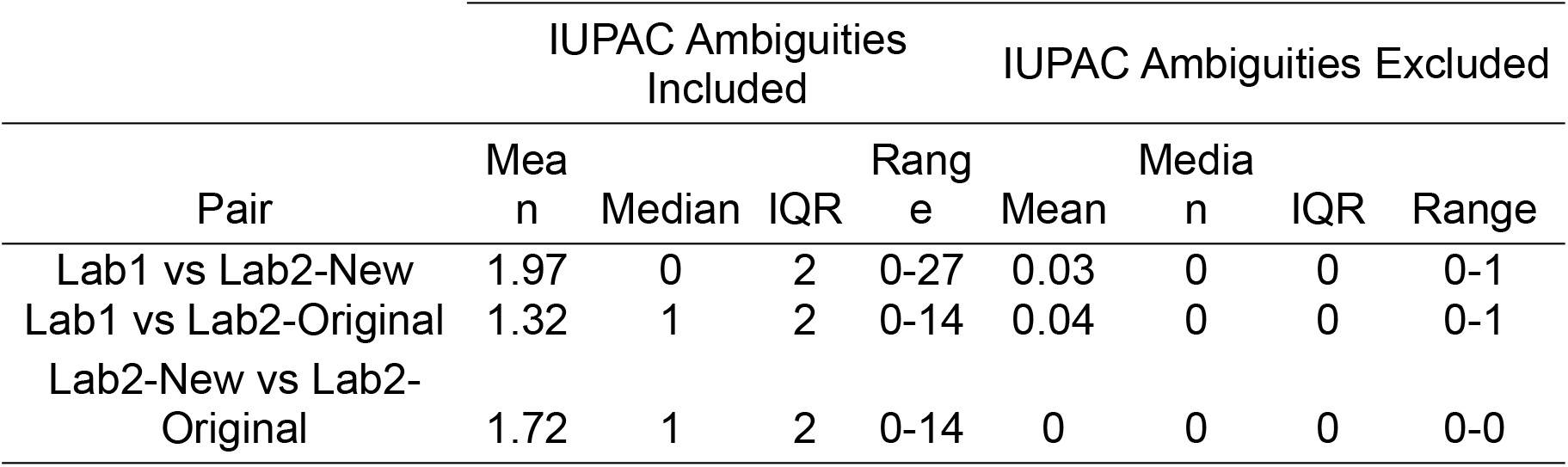
Summary statistics comparing the number of differences in pairwise sequence comparisons between matched samples from the same patient that were sequenced at different lab sites with/without using the same bioinformatics pipeline. Abbreviations: IQR = interquartile range; Lab1: Lab1 sequence assembled with Lab1 bioinformatics pipeline; Lab2-New: Lab2 sequence assembled with Lab1 bioinformatics pipeline; Lab2-Original: Lab2 sequence assembled with Lab2 bioinformatics pipeline.

### Impact on Pango lineage assignment

Out of all 216 pairwise comparisons, the two lineages being compared were assigned the same Pango lineage in 212 cases (Supplementary Table S2). The only differences arose in (a) Sample 34, where the Lab2-New sequence (B.1.2) was assigned a different lineage to both Lab1 and Lab2-Original (B.1.596), and (b) Sample 39, where the Lab1 sequence (B.1.1) was assigned a different lineage to both Lab2-New and Lab2-Original (B.1.1.70). Notably, the only differences among these sequences occurred at sites where either sequence had an IUPAC ambiguity code, demonstrating that incorporation of IUPAC ambiguities or not can impact Pango lineage assignment. However, for the most part differences among matched sequences had no significant impact on Pango lineage assignment, which is crucial for ongoing genomic surveillance efforts.

### Impact on placement in phylogenetic tree

A maximum-likelihood tree for all samples was inferred, with support for nodes estimated through ultrafast bootstraps (UFboot). Despite the relatively low sequence divergence within the alignment, many nodes within the tree had moderate (UFboot 80–94%) to strong (UFboot ≥95%) support (Supplementary Figure S1). We then assessed whether all three samples associated with a given patient formed a triplet clade within the tree (see Methods). Out of 72 possible matched-sample triplets, 62 (86%) formed triplet clades in the phylogenetic tree. Each of these triplet clades received strong UFboot support (range: 97–100%; median: 100%) (Supplementary Table S3). Those samples that did not form triplet clades were generally within slightly larger clades of ∼6 sequences (range: 4-15, Supplementary Table S3), representing a lack of divergence between patients with very similar SARS-CoV-2 sequences. Repeating the analyses using an alignment with problematic sites masked (see Methods) largely did not change the results. That is, the same triplet clades from the unmasked alignment were recovered, with the exception of one poorly supported node for one sample (results not presented). Overall, differences between matches samples did not impact their expected placement within the phylogenetic tree in most cases.

## Discussion

Molecular epidemiology is vital for investigating the origin of SARS-CoV-2, and tracking its spread during the COVID-19 pandemic. An obvious requirement of molecular epidemiological techniques is that inferences are accurate, and that any uncertainty is adequately reported. Most assessments of the reliability and accuracy of molecular epidemiological inference in SARS-CoV-2 research focuses on phylogenetic analysis of SARS-CoV-2 genomes, such as the impact of masking problematic sites on phylogenetic inference (15). Here, we investigated the impact of differences in processing and assembly of raw data for consensus genome generation, which has obvious impacts on downstream bioinformatics analyses. Whilst this study focused on the processing of sequencing reads, any differences in wet lab protocols prior to sequencing may also introduce biases that can propagate through to the stage of bioinformatic analyses.

The extent of differences between matched sequences depended on whether sites with IUPAC ambiguity codes were included in analyses. When these ambiguous sites were excluded, we observed no differences between matched samples in the majority of comparisons. If the bases in the consensus genomes had been purely assigned based on the most common SNV at a given site in the genome, rather than as a function of the frequency of different SNVs at a given position (see Methods), the resulting consensus genomes for matched samples across different laboratories would have been essentially 100% concordant. However, when including ambiguous sites, comparisons between sequences from Lab1 and Lab2 revealed at least one difference between matched samples in 53% of the pairwise comparisons, even when both sequences were assembled using the same bioinformatics pipeline (Table1; Supplementary Table S2).

In a non-haploid organism, IUPAC ambiguity codes are used to represent heterozygous positions in a consensus contig. Given that SARS-CoV-2 is a positive-sense single-stranded RNA virus (2), nucleotide diversity at a particular genomic site does not demonstrate that a virus is heterozygous at that site. Although some sequencing errors are inevitable, nucleotide diversity at a given genomic site more likely represents intra-host diversity (19, 20). Possible causes for intra-host diversity are a given host being infected by SARS-CoV-2 multiple times, or mutations accumulating within different copies of the SARS-CoV-2 genome throughout the course of infection of an individual.

The number of differences in each sample relative to the SARS-CoV-2 reference genome was expected to differ between samples, based on date of collection, and time within the host. Some samples were collected early in the COVID-19 pandemic (March 2020), whereas others were collected many months later (up to the end of November 2020). Accordingly, samples with a greater number of mutations relative to the reference genome also had a greater number of sites at which intra-host diversity could be observed.

There is no threshold beyond which the number of ambiguous bases becomes problematic. However, in practice, any samples that have large numbers of ambiguous sites can be flagged for further investigation and review. For example, an overabundance of sites in a given genome with a roughly 50/50 split of alternate nucleotides can indicate a mixed sample. Our results suggest that most matched samples in our study were highly similar, with a small number of sequences contributing to elevated pairwise differences among matched samples (Table 1). Closer inspection of the sample with the greatest number of differences between laboratories demonstrated extensive intra-host diversity (Sample6, 27 differences, Supplementary Table S2). The majority of differences between laboratories were due to variants occurring at a very low frequency that falls close to our threshold for variant allele frequency, suggesting that only minor differences in sequencing could cause these minor variants to be detected or missed. However, one difference between Lab1 and Lab2 occurs at a non-ambiguous site, and at some positions there are variants at moderate variant allele frequencies (∼0.25-0.5) in the sample from one laboratory, but with no support for a variant in the corresponding sample from the other lab (results not presented). Therefore, ‘Sample6’ from Lab1 might not actually be from the same patient as ‘Sample6’ from Lab2. This investigation demonstrates that (a) incorporating IUPAC ambiguities into consensus genomes is beneficial for quality control, but also that (b) a strict variant allele frequency threshold for IUPAC ambiguity code incorporation (e.g., 0.9) might disproportionately inflate the apparent difference between samples at the consensus level when many minor variants occur at a very low frequency.

Not all bioinformatic software handle ambiguous bases in the same way (34). The tool for Pango lineage designation for molecular epidemiological inference of SARS-CoV-2 genomes is *pangolin* (https://github.com/cov-lineages/pangolin). *Pangolin* can assign lineages using the pangoLEARN machine learning algorithm. Currently, sites with ambiguous data (either ‘N’ or IUPAC ambiguity codes) are ignored by *pangolin* when assigning lineages using the decision-tree algorithm of pangoLEARN. Versions of *pangolin* ≥3.0 can also designate lineages to samples using an ultrafast, parsimony-based tool to place samples in a phylogenetic tree (UShER) (35). The UShER tool replaces ambiguous sites with any one of the most parsimonious nucleotides at that position giving preference, when possible, given to the reference base. Version of *pangolin* ≥3.0 can identify lineages as variants of concern by considering diagnostic constellations of SNVs (scorpio, https://github.com/cov-lineages/scorpio). If there is an ambiguous base at a site in the SARS-CoV-2 genome that is key for assigning lineages, it is possible that an incorrect lineage will be assigned by either skipping that key site (pangoLEARN), or falsely assigning the reference base to that site (UShER). Typing lineages using scorpio should be relatively more robust than strictly using pangoLEARN or UShER, but the method (a) could still fail if IUPAC ambiguities are found at multiple diagnostic SNV locations, and (b) is restricted to designating a set of variants of concern.

Accordingly, we expected that observed differences in the content of ambiguous sites between the sequences of matched samples would result in different Pango lineage assignments, especially when many such differences were observed. However, encouragingly, all but four pairwise comparisons between matched samples revealed an identical lineage assignment. This result most likely occurred because the sites that differ between samples occur in positions of the SARS-CoV-2 genome that are not important for Pango lineage assignment at this stage of the COVID-19 pandemic. As the COVID-19 pandemic continues and more sites become diagnostic for different Pango lineages, the potential issue of base ambiguity will likely become more pronounced if relying on the decision tree approach of pangoLEARN.

Another common use of SARS-CoV-2 genomes is to infer the relationships among samples using a phylogenetic tree. Different phylogenetics software account for IUPAC ambiguity codes in different ways: BEAST, one of the most commonly used Bayesian phylogenetic inference software packages, treats IUPAC ambiguity codes as missing data by default (36), whereas the popular maximum-likelihood program IQTREE accounts for IUPAC ambiguities (33). Therefore, if these programs are used naively, there is a possibility that differences in inferred trees could be caused by IUPAC ambiguity content, either by conflicting phylogenetic signal or a loss of phylogenetic signal. Overall, the extent to which IUPAC ambiguities affect inferred phylogenies is unclear, with conflicting results depending on the taxonomic scale of study organisms and the type of genetic data (34, 37).

The impact of inferred ambiguity codes on placing SARS-CoV-2 sequences in a phylogeny was assessed by estimating a phylogenetic tree using IQTREE2. Three matched consensus sequences were included per sample (see Methods), for a total of 216 sequences. If IUPAC ambiguity codes had a negligible impact on phylogenetic inferences, all matched sequences from a given RNA extract should form a monophyletic clade of three sequences (‘triplet clades’). The vast majority of matched samples did form strongly supported triplet clades (Supplementary Table S3), and those that were not recovered in triplet clades were in regions of the tree with poor support. Any differences in placement could also have been caused by relatively large amounts of missing data originating from amplicon drop out during sequencing. Therefore, any site-by-site differences in sequence content between matched samples, which largely comprise IUPAC ambiguity codes, generally did not affect the placement of a given sample in a phylogenetic tree. In general, these results are consistent with molecular epidemiological inference not being affected by which laboratory a sample is sequenced in.

Overall, despite recovering some differences between matched samples, these differences constituted only a small fraction of the SARS-CoV-2 genomes. Furthermore, these small differences had no impact on Pango lineage assignment or on placement of samples in a phylogenetic tree in the vast majority of cases. These findings demonstrate that: (a) SARS-CoV-2 nucleic acid extracts are relatively robust to transport on dry ice between laboratories, given proper handling, which is promising for laboratories that have the capacity to test for SARS-CoV-2 positivity, but do not have WGS capacity; and (b) any minor differences between sequencing laboratories will likely have a negligible impact on molecular epidemiological inferences based on Pango lineage calls or placement in a phylogenetic tree. However, most of the differences recovered between sequences are only apparent because of our use of a bioinformatics pipeline with strict consensus genome assembly parameters leading to IUPAC ambiguity codes within genomes. Any molecular epidemiological clustering analyses based on pure SNV differences between samples will be vastly different if genomic sites with IUPAC ambiguities are included and different sequencing facilities use different consensus generation thresholds. Therefore, justification and reporting of the parameters used during consensus genome assembly is important, and depends on the intended purpose of the consensus sequence. For example, is the intention to have the sequence represent the most common base at each position in the genome, irrespective of any variation? Or is the sequence intended to represent all variation within a sample, which, in the case of SARS-CoV-2, likely reflects intra-host variation? The latter approach is arguably more biologically realistic and conveys more information, assuming true heterogeneity is represented rather than methodological artifacts, but results in a higher proportion of ambiguous bases in the consensus genome. Overall, we showed that a small number of differences among consensus genomes of matched samples could occur, even when they were assembled using the same bioinformatic pipeline. Careful use of appropriate, comparable pipelines allows appropriate assignment of lineage for use in public health responses.

## Data Availability

The raw sequencing reads from Lab1 are available from the Sequence Reads Archive (SRA) under accession number PRJNA750251. GISAID and SRA accessions for Lab2 are given in Supplementary Table S1. The custom python3 program for pairwise comparisons is available from https://github.com/charlesfoster/pairwise_comparisons. The custom R script for comparing sequence triplicates is available from https://github.com/charlesfoster/useful_scripts/blob/master/find_sample_triplets.R.

## Acknowledgements

We thank the staff within NSW Health Pathology Diagnostic laboratories at the Serology and Virology Division (SAViD), including Anna Condylios, Dr Zin Naing, Jane Shi, and Elsa Liu; staff of the NSW Health Pathology-Institute of Clinical Pathology and Medical Research, including Drs Jen Kok, Mailie Gall, Jenny Draper, Elena Martinez, Clement Lee, Qinning Wang, and Andrew Ginn; and staff of the Centre for Infectious Diseases and Microbiology-Public Health, Westmead Hospital, including Connie Lam, Verlaine Timms, Rosemarie Sadsad, and Karen-Ann Grey. All staff acknowledged here contributed to data generation.

## Table and Figure Legends

**Supplementary Figure S1:** Phylogenetic tree inferred using maximum likelihood analysis with IQTREE2. Support for nodes is represented with ultrafast bootstrap support; scale bar is in substitutions per site. Abbreviations: samples ending with “_L1” = primary assembly from Lab1, samples ending with “_L2-O” = primary assembly from Lab2, samples ending with “_L2-N” = secondary assembly of sequencing reads from Lab2.

**Supplementary Table S1:** Accessions for all sequences sourced from GISAID, as well as the sample numbers designated in this study.

**Supplementary Table S2:** Results of pairwise comparisons between matched samples, with and without ambiguous sites included in the analysis. Pango lineages were inferred using the pangoLEARN algorithm of *pangolin* v2.3.6.

**Supplementary Table S3:** Table depicting whether all three sequences from matched samples formed a monophyletic clade of three tips (‘triplet clade’), as well as ultrafast bootstrap node support for the clade.

