## Supplementary figures and images for "Assessment of inter-laboratory differences in SARS-CoV-2 consensus genome assemblies between public health laboratories in Australia"

### Supplementary Figure S1

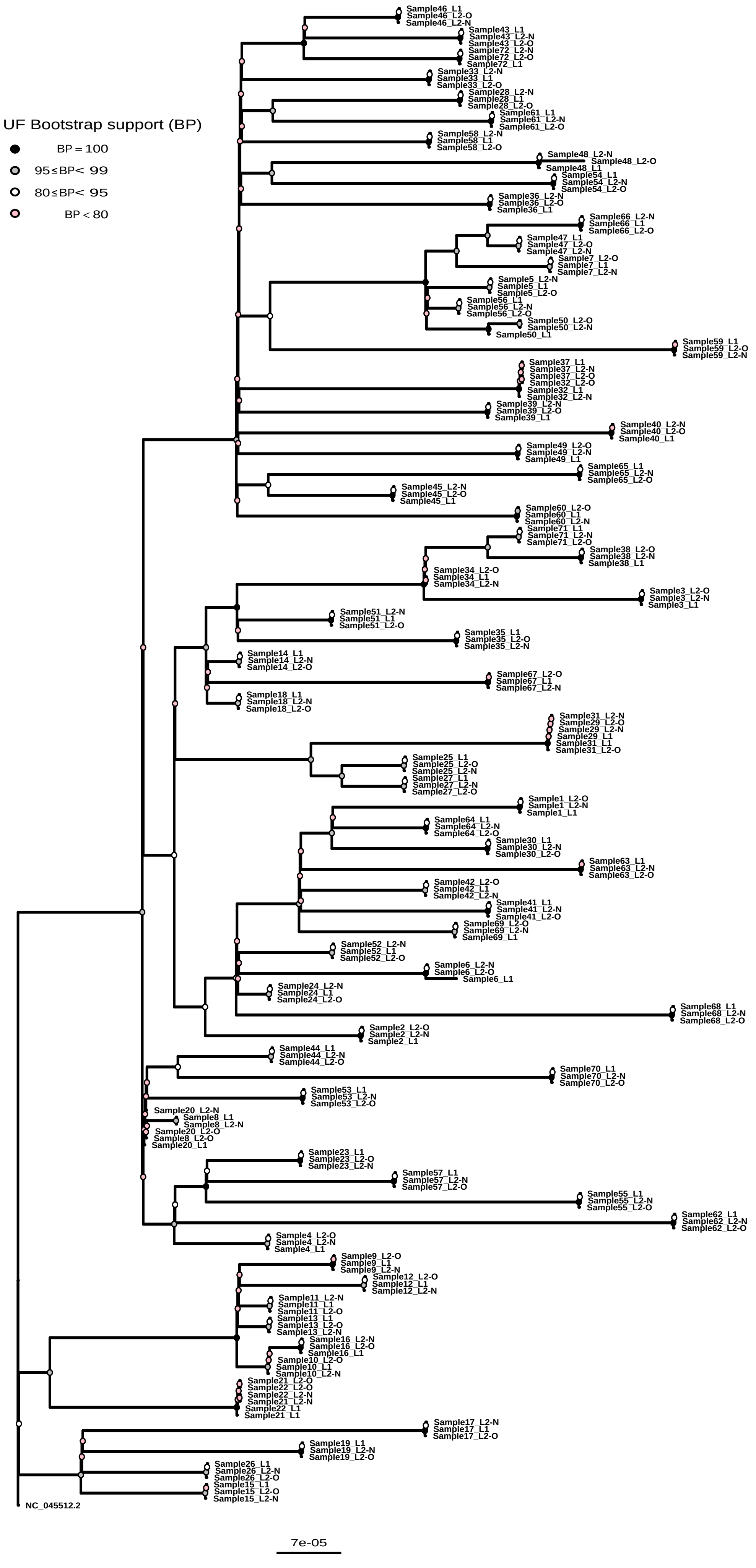
